# Birhan Maternal and Child Health cohort: a study protocol

**DOI:** 10.1101/2021.01.29.21250334

**Authors:** Grace J. Chan, Bezawit Mesfin Hunegnaw, Kimiko Van Wickle, Yahya Mohammed, Mesfin Hunegnaw, Frederick G. B. Goddard, Fisseha Tadesse, Delayehu Bekele

## Abstract

**Introduction:** Reliable estimates on maternal and child morbidity and mortality are essential for health programs and policies. Data are needed in populations which have the highest burden of disease but also have the least evidence and research, to design and evaluate health interventions to prevent illnesses and deaths that occur worldwide each year.

**Methods and analysis:** The Birhan Maternal and Child Health (MCH) cohort is an open prospective pregnancy and birth cohort nested within the Birhan health and demographic surveillance system (HDSS). An estimated 2500 pregnant women are enrolled each year and followed through pregnancy, birth, and the postpartum period. Newborns are followed through two years of life to assess growth and development. Baseline medical data, signs and symptoms, laboratory test results, anthropometrics, and pregnancy and birth outcomes (stillbirth, preterm birth, low birthweight) are collected from both home and health facility visits. We will calculate the period prevalence and incidence of primary morbidity and mortality outcomes.

**Ethics and Dissemination:** The cohort has received ethical approval. Findings will be disseminated at scientific conferences, peer-reviewed journals, and to relevant stakeholders including the ministry of health.

**Strengths and limitations of this study:**

- This cohort collects longitudinal data at multiple time points from pregnancy through birth and childhood in a setting where there are limited data.
- Data from this study provide estimates for birth outcomes such as stillbirths, preterm birth, and low birth weight.
- Results will inform risk profiles for maternal, neonatal, and child morbidity and mortality.
- Similar to all observational studies, there are potential confounders that are unmeasurable.
- Evidence from this study will support policies and programs to improve maternal and child health.

## INTRODUCTION

Globally, there has been significant progress in improving maternal and child health over the past few years. However, maternal, neonatal, and child mortality remains disproportionately high in parts of the world that have limited data, research, and programs. Almost 94% of maternal mortality occurs in low-middle income country (LMIC) settings from preventable causes.^1^ Sub-Saharan Africa has the highest under-five mortality.^2^ There is a significant gap in our knowledge of the rates of stillbirths and where they occur. Globally, an estimated 2.6 million (uncertainty range 2.4-3.0 million) third trimester stillbirths occurred in 2015 with 98% estimated to have occurred in LMIC settings.^3 4^

In Ethiopia, the second most populous country in Africa, the maternal mortality rate is 412/100,000 live births totaling 14,000 maternal deaths each year.^5-7^ Ethiopia is one of five countries which contributed to more than half of the global under-five deaths in 2018.^8^ With 80,000 deaths each year, Ethiopia is one of ten countries accounting for more than half of global neonatal deaths.^9^ The rate of stillbirths at a population-level in Ethiopia remains unmeasured. Rough estimates range from 9.2 stillbirths per 1000 births based on data from the 2016 Ethiopia Demographic and Health Survey (EHDS) maternal report of a pregnancy loss after seven months gestation in the last 5 years to 28 stillbirths per 1000 births in 2012 from a systematic review of predominantly hospital-based studies.^5 10 11^

Priority-setting for policies, programs and research to prevent adverse maternal and neonatal outcomes relies on accurate and timely health data.^12^ Data are needed for the implementation of health interventions to prevent maternal and child deaths and morbidities. However, data on basic vital statistics is lacking in most low-income countries. In Sub-Saharan Africa, for example, fewer than ten countries have vital registration systems that produce usable data.^13^

There are significant gaps in counting every pregnancy, newborn, and child; identifying high-risk populations; and understanding causal pathways for morbidity and mortality to develop and test interventions. Many maternal and neonatal deaths are preventable with improved identification of high-risk pregnancies; treatment and management of maternal complications; and improved quality of care at birth and during the postnatal and early childhood period.^3 4^ Access to timely, accurate, and dependable information is essential to improving health and health systems. To address these gaps, the Birhan Maternal and Child (MCH) cohort is designed to describe the epidemiology of maternal and childhood health outcomes and risk factors in North Shewa Zone, Amhara Region, Ethiopia. Specific objectives include:

1. To estimate the incidence of birth outcomes, including stillbirths, early neonatal mortality, preterm birth, and low birth weight.
2. To identify severe illnesses and deaths among pregnant women and children under-2 years old and identify risk factors for morbidity and mortality.
3. To build the capacity of the Ethiopian Federal Ministry of Health (FMoH) and local institutions to generate and use local evidence for action to improve maternal and child health.

## METHODS AND ANALYSIS

### Study design

The Birhan MCH cohort is an open prospective pregnancy and birth cohort nested within the Birhan health and demographic surveillance system (HDSS). Briefly, the Birhan HDSS includes 16 kebeles (lowest administrative unit) in two districts and as of July 2019 includes a total of 18,933 households and 77,766 people.^14^ Basic health and demographic data are collected every three months through house-to-house surveillance. During these quarterly visits, data collectors conduct pregnancy surveillance among married women of childbearing age through a series of pregnancy screening questions. Urine pregnancy tests are done for women who screened positive. Pregnant women who consent to participate are enrolled in to the MCH cohort. Mothers of and children under two are also enrolled into the MCH cohort. Both clinical and epidemiological data are collected at health facilities during antenatal, postnatal, and sick visits and in the community during scheduled home visits.

### Study population

Pregnant women, mothers of children under two years of age, and children under-two from the Birhan catchment population are eligible for enrollment if they meet the following inclusion criteria:

- Provide informed consent.
- Enrolled in the Birhan health and demographic surveillance system. Pregnant women must have a confirmed pregnancy (urine pregnancy test, ultrasound, or other positive signs of pregnancy).

Women and children are excluded from the study if they are from:

⍰ Semi-formed populations living in the camps e.g., military personnel and prisoners.
⍰ Street children/orphans who have no address as they are difficult to follow
⍰ Those who come to the area only for work during the day and have a primary household outside the catchment area

### Follow-up schedule

Once enrolled, pregnant women are followed both at home and at the health facility. The home visits are conducted every three months until 32 weeks of gestation, then every two weeks. After 36 weeks, the home visits are alternated with a phone visit every week until birth. Postpartum women are followed from birth to 42 days postpartum with scheduled home visits on days 0 (birth), 6, 28, and 42 after birth. Newborns and children under-two years are followed on days 0 (birth), 6, 28 and 42 and months 6, 12, and 24. Participants who present to health facilities for antenatal care, postnatal visits, or sick visits (outpatient or admission) also have data collected by study data collectors at the health facility. Participants are censored from the study with the following events: out-migration from the catchment area, death, lost to follow-up, and at the end of the follow-up period (See Figure 1). All participants are concurrently followed on a quarterly basis by the underlying HDSS.

**Figure 1.**
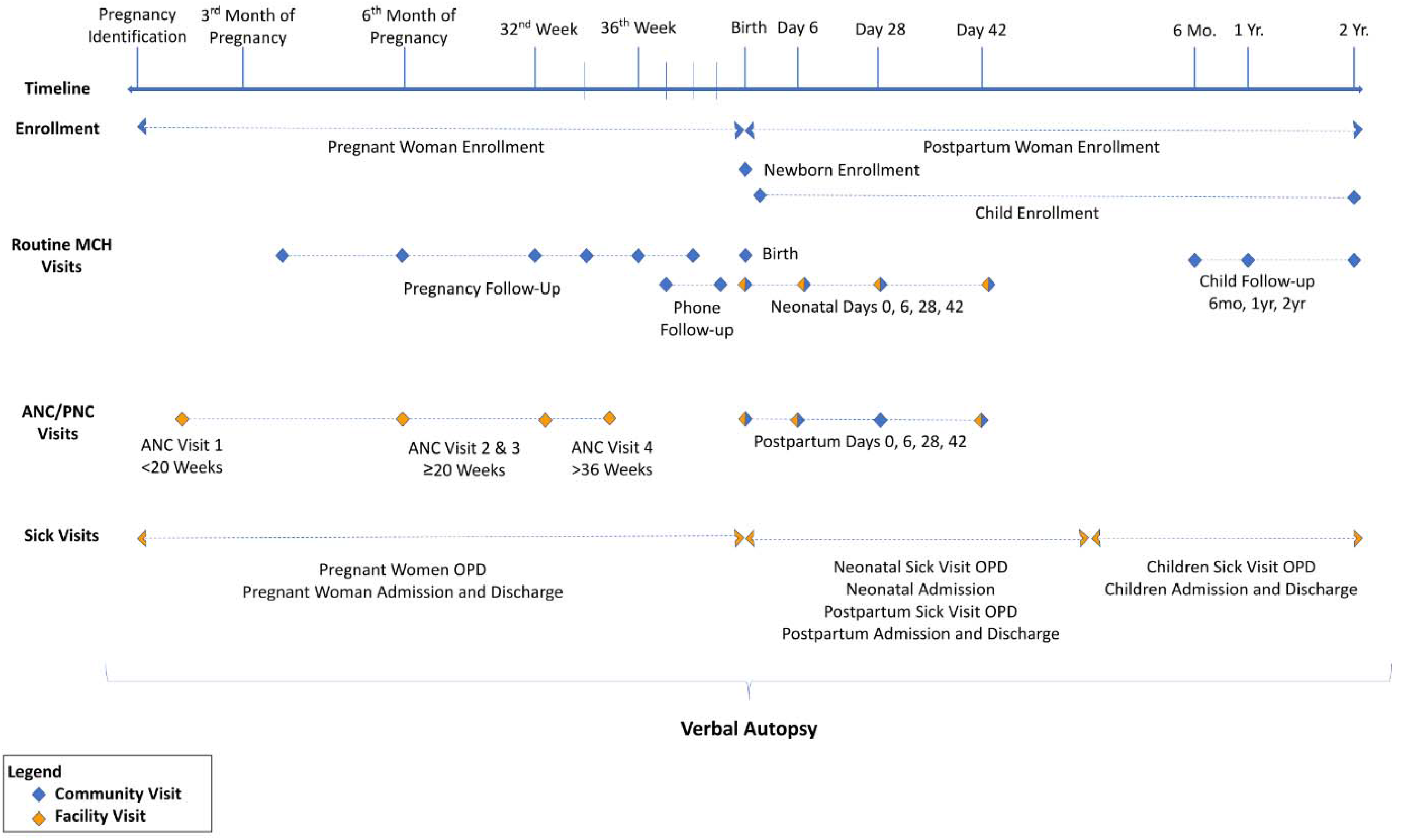
Schedule of study enrollment and follow-up visits

### Data collection

Data are collected by several sources: maternal or caretaker recall, data collector assessments in the community, health worker observations at the community, health center, and hospital level. Data collected at the time of HDSS enrollment include household data on socioeconomic status, drinking water and sanitation access, flooring/wall/roof materials, number of rooms used for sleeping, place of cooking, household possessions and ownership of land. Individual (mother, father, child, grandparent) specific data on education, employment, literacy, religion, care seeking behaviors, and immunization rates are collected. Anthropometric data including weight, height, and mid-upper arm circumference (MUAC) measurements are collected for women of child-bearing age and children under-two.

Pregnant women enrolled into the cohort receive an ultrasound for gestational age dating at their earliest ANC visit. Information on the pregnancy (birth spacing, antenatal care visits) and pregnancy-related symptoms, including history of vaginal bleeding, dysuria, headache, fever, abdominal pain, and fetal movement are collected. Maternal weight and observations of clinical signs such as pallor, jaundice, and edema are collected. For newborns enrolled in the cohort within 24 hours of birth, data on labor and delivery (location of delivery, delivery method, duration of labor, complications of birth, birth weight) and immediate newborn care (breastfeeding, cord-care, bathing, skin emollients) are collected. Mothers or caretakers recall of clinical symptoms, observations of signs of illness, and neonatal weight are collected at subsequent postpartum visits. Clinical symptoms, observations of signs of illness and possible risk factors such as sick contacts, food sources, travel, and water sanitation and hygiene practices are collected at sick visits.

To ascertain the cause of death, verbal autopsies are conducted to obtain information on the symptoms, signs, and other relevant events during the illness leading to death. Three verbal autopsy questionnaires (for deaths 0-28 completed days of life; deaths of children between four weeks and 11 years of age; and deaths of persons aged 12 years and above) adopted from the standard 2016 WHO VA questionnaires are used. ^15^ Verbal autopsies were developed to provide information on the cause of death in communities where there is limited access to health care and medical certification of causes of death. In such situations the only viable source of information on the terminal illness is from caregivers of the deceased, most often family members. Ascertaining causes of death from such information is based on the premise that VA respondents can accurately recall the details of the various symptoms and events that occurred during the period of illness prior to death, and that such information can be used to classify the cause(s) of death into diagnostic categories based on specific symptom complexes.

### Electronic data collection system and data management

We use the Birhan electronic data collection system, built from Open Data Kit (ODK) ^16^, for longitudinal and relational data collection. ODK is a free, open-source application used to facilitate mobile data capture. ODK can be coded using SQL to facilitate data collection, transfer and storage, and authoring of the electronic questionnaires used in the data collection process. Data can be collected on a mobile device off-line. Data are uploaded daily using mobile data to the central database. Data quality checks and logic are built into the data entry tools and data system. Data are hosted on an encrypted central database on the cloud and are de-identified. The data system is developed and maintained by two data system developers. Data are managed by a team of data scientists and data managers in Stata (Version 15.1) and R/RStudio (Version 3.6.2).

### Data quality

To ensure high data quality, we developed simple user-friendly questionnaires which were piloted prior to use. We recruited and trained data collectors who met a minimum level of competency through examinations. Supervisors oversee study implementation by supporting data collectors with weekly on-the site mentorship and weekly team meetings. Supervisors independently conduct home visits on a 5% random sample of households to validate data collector performance. Within the electronic data capture system, we built single field value checks, inter-field logic checks and inter-form logic checks. In case of an error at the time of data entry, pop-up warnings are triggered, prompting the user to resolve any issues prior to saving the record. Following data collection, the dataset is systematically checked for data quality issues by a team of analysts. Data quality issues are recorded, provided to data collectors to identify solutions, and then rectified in the dataset.

### Primary outcomes

Primary outcomes include:

- Severe maternal morbidity and maternal mortality: unintended outcomes of the process of labor and delivery that result in significant short- or long-term consequences to a woman’s health
- Stillbirth: death prior to delivery of a fetus ≥28 weeks of gestation (or >1000 grams if gestational age is unavailable)
- 7-day neonatal mortality: death of a live-born infant at 0-6 days of life
- 28-day neonatal mortality: death of a live-born infant at 0-27 days of life
- Preterm birth: delivery prior to 37 completed weeks of gestation of a live born infant
- Low birth weight: birth weight <2500 grams of live-born infant
- Place of birth (home versus facility)
- Child morbidity: incidence of diarrhea, pneumonia, febrile illnesses^17 18^
- Causes of mortality: Numbers and rates of maternal, childhood, and adult deaths and causes of death by verbal autopsy.^19 20^

Secondary outcomes include recurrence of illnesses, readmissions, and growth as measured by weight, height, MUAC, and body mass index.

### Data analysis

For each of the primary outcomes, we will calculate the period prevalence and incidence at community and health facilities levels. To estimate period prevalence, we will sum the two-week periods over all the participants within certain age ranges as the denominator and sum the number of episodes within that two-week period as the numerator (i.e., cases over sum of person-days observed or person-days recalled). We will estimate the incidence for each outcome by taking the sum of all the person-time contributed by each person as the denominator, and the sum of all episodes over the study period as the numerator. Missing data will be assessed through quality assurance checks by field supervisors and rectified through a documented error correction system. Remaining incomplete data will be addressed through analytical approaches. We will repeat the above analyses with varying case definitions of disease severity.

In a population of 77,000 we expect approximately 2500 pregnancies per year and 4500 children under-2 years of age. Using historical data and the literature, we expect a range of outcome rates from 2.0% to 20.0% depending on outcome of interest (Table 1). The prevalence of stillbirths is estimated to be 9.2 per 1000 births in Ethiopia and 19.7 per 1000 births (around 2.0%) in Amhara.^21^ The prevalence of diarrhea over a two-week period among children under-two varies by region: 31.3%^22^ in Afar, 18.5% in SNNP, 16.0% in Oromia, 17.7% in Amhara, 6.8% in Tigray^23^ Given this variation, we estimate the prevalence of diarrhea over a two-week period among children under-two to be 20.0%. We expect the prevalence of acute respiratory infection over a two-week period among children under-two to be 5.0% based on preliminary data in Ethiopia.^23^

**Table 1.** Outcomes of interest, estimated prevalence, and precision (95% CI width) by sample size∗

| Outcomes | Estimated prevalence | 2500 | 3500 | 4500 |
| --- | --- | --- | --- | --- |
| Stillbirth<sup>21</sup> | 2.0% | 0.6% | 0.5% | 0.4% |
| Neonatal death (28-day)<sup>5</sup> | 3.0% | 0.7% | 0.6% | 0.5% |
| Acute respiratory infections** | 12.0% | 1.3% | 1.1% | 0.9% |
| Diarrhea**<sup>23</sup> | 20.0% | 1.6% | 1.3% | 1.2% |

\* Precision calculated using exact distribution for <5% prevalence and Wald distribution for ≥5% prevalence
\*\*Estimated two-week point prevalence for children under-2 years old

To estimate absolute precision (defined as the half-width of the 95% confidence interval) for outcomes ranging in prevalence from 2.0% to 20.0%, the exact (Clopper Pearson) distribution was used for rare outcomes (<5%) and the Wald distribution was used for common outcomes (≥5%).^24 25^ A sample size of 2500 pregnant women would estimate the prevalence for a rare outcome such as stillbirth, 2.0%, with 0.6% precision (95% CI 1.4% to 2.6%) (Table 1). For more common outcomes such as preterm birth, a sample size of 2500 pregnant women would estimate a 12.0% prevalence with 1.3% precision (95% CI 10.7% to 13.3%). A sample size of 4500 children would estimate a 20.0% prevalence of diarrhea with 1.2% precision (95% CI 18.8-21.2%). A sample size of 4500 children would estimate a 5.0% prevalence of ARI with 0.5% precision (95% CI 4.5-5.5%). Period prevalence will vary based on case definitions and severity of illnesses. To examine risk factors and correlates for these outcomes, we will conduct tests of association to detect statistically significant effect sizes for risk factors and outcomes assessed in this study.

## Data Availability

Data use is governed by the Birhan Data Access Committee (DAC) and follows Birhan's data sharing policy. All researchers who wish to access Birhan data can complete a Birhan data request form and submit it for decision by the Birhan DAC. Datasets will only be provided with de-identified data to maintain confidentiality of study participants.

## ETHICS AND DISSEMINATION

The study has received the following ethical approvals: St. Paul’s Hospital Millennium Medical College (PM23/274), Boston Children’s Hospital (P00028224) and Harvard School of Public Health (19-0991). Results will be made available to the ministry of health, regional, zonal and district health offices. Publications will follow the HaSET publications guidelines. Birhan serves as the field site for HaSET (‘happiness’ in Amharic)—a maternal and child health research program in Ethiopia. For further details, please visit www.hasetmch.org.

### Patient and public involvement

The scope of the research questions and outcome measurements were informed by identified gaps in maternal and child health and key priorities at the Ministiry of Health including limited reliable health and demographic data and maternal and child health outcomes, that is needed to support evidence-based decision making. To build capacity on research methodologies, data analyses, and interpretation of data, fellows and trainees are involved with the cohort study. A community advisory board is engaged with the research.

## ACKNOWLEDGEMENTS

We are grateful to the community of the Birhan field site, especially the mothers and children who participate. We appreciate the generous support from the Ministry of Health, Amhara Regional Health Bureau, North Shewa Zone Office, Angolela Tera and Kewet/Shewa Robit Woreda Health Offices and catchment health facilities. We thank the data collectors, supervisors, coordinators and the HaSET team for their contributions to the Birhan MCH study. St. Paul’s Hospital

## CONTRIBUTORS

The study concept was conceived by GC and DB. GC, DB and BH contributed with study design and implementation. MH and CB led the data collection. YM, KW, FG contributed to data management. Analysis will be performed by KM, KW, and FG. GC drafted the initial manuscript. All authors contributed to the refinement of the protocol, provided edits, and critiqued the manuscript for intellectual content.

## COMPETING INTERESTS STATEMENT

None declared

## FUNDING

This work has been supported by the Bill & Melinda Gates Foundation (Grant number INV-010382 and OPP1201842).

## Notes

### Competing Interest Statement

The authors have declared no competing interest.

### Author Declarations

The study has received the following ethical approvals: St. Paul's Hospital Millennium Medical College (PM23/274), Boston Children's Hospital (P00028224) and Harvard School of Public Health (19-0991). Results will be made available to the ministry of health, regional, zonal and district health offices. Publications will follow the HaSET publications guidelines. Birhan serves as the field site for HaSET ('happiness' in Amharic)-a maternal and child health research program in Ethiopia. For further details, please visit www.hasetmch.org.

